# Legumes as a substitute for red and processed meat, poultry, or fish, and the risk of non-alcoholic fatty liver disease in a large cohort

**DOI:** 10.1101/2024.12.09.24315856

**Authors:** Fie Langmann, Daniel B. Ibsen, Luke W. Johnston, Aurora Perez-Cornago, Christina C. Dahm

**Affiliations:** Aarhus University, Department of Public Health, Bartholins Allé 2, 8000 Aarhus C, Denmark; Steno Diabetes Center Aarhus, Aarhus University Hospital, Palle Juul-Jensens Boulevard 11, 8200 Aarhus C, Denmark; European Commission, Joint Research Centre (JRC), Ispra, Italy

**Keywords:** dietary pulses, legumes, non-alcoholic fatty liver disease, NAFLD, nutritional epidemiology, dietary substitution model

## Abstract

**Background:** Dietary recommendations have globally shifted towards promoting consumption of legumes as an environmentally friendly and healthy source of protein. This study investigated replacement of red and processed meat, poultry, or fish for equal amounts of legumes on the risk of non-alcoholic fatty liver disease (NAFLD).

**Methods:** UK Biobank participants who completed ≥2 dietary assessments and had complete covariate information were included in the analyses (N=124,194). Information on dietary intake was collected using two to five 24-hour dietary assessments. Incident cases of NAFLD were determined through linkage to the National Health Service registries. The rate of developing NAFLD when replacing 80 g/week of red and processed meat, poultry, or fish with legumes was estimated using multivariable-adjusted Cox proportional hazards regression.

**Results:** During follow-up (median 10.49, IQR: 10.4-10.9 years), 1201 individuals developed NAFLD. Replacing 80 g/week red and processed meat or poultry with legumes was associated with 4% and 3% lower rates of NAFLD, respectively (meat HR: 0.96, 95% CI: 0.94; 0.98; poultry HR: 0.97, 95% CI: 0.95; 0.99). Replacing 80 g/week of fish with legumes was not associated with NAFLD (fish HR: 0.98, 95% CI: 0.96; 1.01). Results did not change markedly after adjustment for BMI.

**Conclusion:** Consuming one serving of legumes weekly instead of red and processed meat or poultry was associated with a slightly lower rate of NAFLD, while consuming legumes instead of fish did not show an association with NAFLD. Further research in cohorts with higher legume consumption is needed to confirm these findings.

**Highlights:** Food substitution models improve interpretation of studies of dietary exposures in observational studies.

Replacing red and processed meat or poultry with legumes was associated with slightly lower rates of non-alcoholic fatty liver disease in the UK Biobank. No association was found when replacing fish with legumes.

## Introduction

Non-alcoholic fatty liver disease (NAFLD), defined as liver fat content of more than 5% without secondary causes of hepatic fat accumulation, is the most prevalent chronic liver disease with a global prevalence of 25%.(^1^) NAFLD is associated with an increased risk of cardiovascular diseases(^1^) and can furthermore progress to non-alcoholic steatohepatitis (NASH), which may further predispose individuals to develop fatal liver complications.^(2; 3)^ Long-term exposure to Western diets, obesity, physical inactivity, and smoking are all risk factors for NAFLD.(^4^) Concurrently, NAFLD and NASH increase the levels of liver enzymes like alanine aminotransferase (ALT), which have been associated with increased incidence of all-cause and cancer mortality.^(5; 6)^

Food production and consumption contribute up to 42% of global CO_2_ emissions.^(7; 8)^ To reduce climate impacts of human diets, sustainable dietary guidelines have been introduced emphasising greater consumption of plant-based protein sources such as legumes.^(9; 10)^ The climate impact of legumes is undoubtedly low,(^7^) however the scientific evidence on the association between legume intake in isolation and health outcomes is sparse.(^11^) Legumes are good sources of protein, low in saturated fat and energy density, and rich in dietary fibre; all components associated with a healthy diet.(^12^) Consumption of diets including legumes has been associated with overall better diet quality and greater health compared to consuming Western diets high in red and processed meat, fats, and sugars in observational studies.(^13–15^) Animal studies have shown that legume consumption up-regulates lipid metabolism in liver cells, thus minimizing the risk of NAFLD by reducing the build-up of fats in the liver.^(16; 17)^ However, research in humans on the association between legume consumption and NAFLD is sparse and inconclusive.^(11; 18; 19)^

When individuals limit their intake of certain food groups, they will increase the intake of certain other food groups, in an otherwise stable diet.(^20^) Legumes are often included in the diet as a source of plant-based protein, and the health benefits of substituting legumes for other protein-rich foods may depend on the substitute as some sources of protein may be better to replace than others.(^19^) Replacing protein from animal sources with protein from plant sources has previously been associated with a substantially lower mortality rate in some studies.(^21^) Furthermore, even low consumption of red and processed meats has been associated with a greater risk of developing NAFLD, while consumption of seafood or poultry has shown mixed directions of associations.(^1–3^) Therefore, this study aimed to investigate the association between replacing red and processed meat, poultry, or fish with legumes and the risk of NAFLD or NASH contingent on potential confounders of the associations.

## Methods

### Study population and setting

The UK Biobank prospective cohort was established in 2006 and invited 9.2 million people to participate in the study.(^22^) The participation rate of 5.5% resulted in approximately 500,000 participants aged 37-73 years in the study. At baseline, participants provided detailed information on sociodemographic, physical, lifestyle, diet, and health-related characteristics via self-completed touch-screen questionnaires and a computer-assisted personal interview.(^22^) Professionally trained staff took physical, anthropometric, and biomedical measures following standardized procedures.(^22^) Participants’ diets were assessed with up to five 24-hour dietary assessments, using the Oxford WebQ,(^23^) which was completed at least once by 210,950 individuals.

For the current study, only participants who completed two or more 24-hour dietary assessments were included in the analysis to capture usual intake of legumes. Participants with missing information on covariates were excluded apart from missing information on physical activity. If participants were missing information in all these categories, they were excluded from the study; if any information was available, the participant was included and individual missing data points on physical activity were coded as unknown.

All participants gave written informed consent to participate in the study. The UK Biobank was approved by the National Information Governance Board for Health and Social Care and the National Health Service (NHS) North West Multicentre Research Ethics Committee (ref 21/NW/0157). This research has been conducted using the UK Biobank Resource under Application Number 81520.

### Assessment of diet

Individuals recruited in the final recruitment period, between April 2009 and September 2010, completed their first Oxford WebQ at the assessment centre at baseline (N = 70,747). Any participant in the UK Biobank cohort who provided a valid email address was invited to complete the Oxford WebQ online on four separate occasions during follow-up.(^23^)

The Oxford WebQ was designed as an internet-based 24-hour dietary assessment tool. The questionnaire comprises a short set of food frequency questions on commonly eaten food groups in the British population assessing diet in the previous 24 hours.(^23^) The Oxford WebQ was validated against interviewer-administered 24-hour dietary recalls and recovery biomarkers.(^24^) The validation showed similar performance for estimating true intakes of protein, potassium, and total sugars when using two or more dietary assessments compared to the more burdensome interviewer-based recall.(^24^) Recently, the Oxford WebQ nutrient calculation was updated to provide more detailed information on nutrient intakes and to incorporate new dietary variables.(^25^)

Consumption of legumes, red and processed meats, poultry, and fish was based on the total weight of food group intakes estimated as the average from participants’ responses in the Oxford WebQs.(^26^) The food groups used in this study are defined in Supplementary Table 1.

### Non-alcoholic fatty liver

The International Classification of Diseases and Related Health Problems 10^th^ edition disease codes (ICD-10) were used to define disease outcomes.(^27^) The entity metabolic dysfunction-associated steatotic liver disease (MASLD) was defined in 2023 as the new nomenclature for NAFLD and NASH. However, as MASLD is not part of the ICD-10^th^ edition upon which the outcomes are defined and diagnosed, we used NAFLD and NASH throughout this manuscript.(^28^) Incident cases of NASH and NAFLD were assessed through linkage to the NHS registers, where diagnosis after hospital admissions were coded according to the ICD- 10.(^27^) Incident cases of NAFLD were diagnosed with ICD-10-code K76.0 at first admission to the hospital while incident cases of NASH were diagnosed with ICD-10-code K75.8.(^29^)

### Covariates

Covariates were chosen *a priori* based on a review of the literature and directed acyclic graphs (Supplementary Figure 1).

Information on covariates included information on average intakes of all other food groups retrieved from the Oxford WebQ (refined cereal, whole grain cereal, dairy products, dietary fats, fruits, vegetables, nuts, mixed dishes, potatoes and tubers, eggs, non-alcoholic beverages, alcoholic beverages, snacks, and sauces and condiments). Other covariates were self-reported at the baseline visit and included information on sex, age, ethnicity, yearly income, educational level, Townsend Deprivation Index (0 indicates that an individual lives in an area with overall mean deprivation, positive values indicate higher material deprivation while negative values indicate relative affluence),(^30^) geographical region of recruitment,(^23^) cohabitation, anthropometry, physical activity, smoking status, history of NAFLD-related diseases, and family history of NAFLD-related diseases. Usual alcohol consumption (g ethanol/day) was estimated as the average of the responses to the Oxford WebQs. Blood samples were drawn at baseline for all participants to measure ALT. When elevated, ALT serves as a proxy measure for increased risk of developing NAFLD.^(5; 6)^

### Statistical analyses

Standard summary statistics were performed to describe the distribution of participants’ baseline characteristics and food group consumption across strata of legume consumption. The legume consumption strata were defined as non-consumers (0 g/week), low intake (tertile 1 of those who consumed any legumes), medium intake (tertile 2 of those who consumed any legumes), and high intake (tertile 3 of those who consumed any legumes). Summary statistics were also computed to describe baseline characteristics and food group consumption across incident NAFLD.

Multivariable adjusted Cox proportional hazards regression models were used to estimate the hazard ratio for NAFLD or NASH based on substituting equal masses of red and processed meat, poultry, or fish with legumes. The NHS defines one serving of legumes and pulses as three heaped tablespoons or 80 g(^31^) and the substitution magnitude was chosen to reflect a weekly substitution where one serving of legumes or pulses (80 g) replaced an equal amount of either red and processed meat, poultry, or fish. Substitutions were modelled using the leave-one-out approach, following this model:

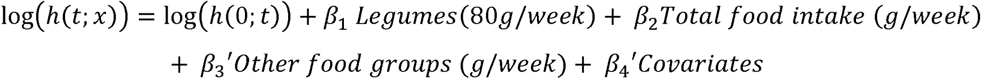

Apostrophes indicate a group of coefficients consisting of several individual variables (β3’ and β4’). Variables for intakes of each food group (β3’) and total food intake (β2) were included and held stable, while the food group that were to be substituted was left out of the model.(^20^) For instance, when estimating the HR for replacing red and processed meat with legumes, 80g of legumes was specified in the model, while the intake of red and processed meat was left out of β3’. Red and processed meat still contributed to the total food intake (β2). The estimated HR thus expressed the association of keeping the total food intake stable while specifying that 80 g/week food should come from legumes instead of red and processed meat.

Age was used as the underlying time scale in the analyses. Person-years at risk were calculated from the date of last completed Oxford WebQ to the date of death, loss to follow-up, diagnosis of NAFLD or NASH, or right censoring, whichever occurred first. As participants in UK Biobank are still followed today, participants were right censored on October 31^st^, 2022, when the most recent registry update of full follow-up for the outcomes was completed. The proportional hazards assumption was evaluated using time-varying covariates and Schoenfeld residuals and revealed no evidence of deviation.

The substitution analyses were conducted with different adjustment levels. Model 1 was stratified by age at recruitment (<45, 45–49, 50–54, 55–59, 60–64, ≥65 years), geographical region of recruitment (ten UK regions),(^23^) and sex, and adjusted for intakes of all other food groups (g/week) apart from the food being replaced. When substituting g/week of legumes for other foods, the standard unit for all dietary elements was also in g/week. This study modelled substitution of foods with varying energy densities.^(32; 33)^ ^(32; 33)^ Nevertheless, to avoid estimating obscure relative effect estimands with unclear interpretations due to different units of measurement in the model, total energy intake was not adjusted for.(^33^) The analyses accounted for all food items based on their weight, and the analyses were adjusted for total amount of food consumed in g/week.(^20^) Model 2 was further adjusted for alcohol consumption (continuous; g ethanol/week as restricted cubic splines with 4 knots), ethnicity (categorical; White, other), educational level (categorical; low: Certificate of Secondary Education (CSE), National Vocational Qualifications, Higher National Diploma, Higher National Certificates, other professional qualifications, or equivalent; intermediate: A levels, O levels, General Certificate of Secondary Education, or equivalent; high: College or University degree), yearly income (categorical; <18,000£, 18,000-30,999£, 31,000-51,999£, 52,000-100,000£, >100,000£, prefer not to answer), Townsend Deprivation Index (continuous), cohabitation (categorical; alone, with spouse or partner, with non-partner, prefer not to answer), physical activity (categorical; low [0-9.9 METs/week], moderate [10- 49.9 METs/week], and high [≥50 METs/week], unknown), smoking status (never, former, current 1-15 cigarettes per day, current ≥15 cigarettes per day, unknown), history of NAFLD-related diseases (categorical; yes, no or don’t know), and family history of NAFLD-related diseases (categorical; yes, no or don’t know). As obesity may either confound or mediate the association between replacing red and processed meat, poultry, or fish with legumes and the risk of NAFLD, model 3 was further adjusted for BMI (categorical; <30 kg/m^2^ or ≥30 kg/m^2^).

### Secondary and sensitivity analyses

Secondary analyses included Cox proportional hazards regression analyses modelled as the main analyses in model 2, but in a study sample restricted to consumers of legumes (i.e. removing non legume consumers). We also modelled legume consumption as a continuous exposure to evaluate the non-substitution association between legume consumption and NAFLD with an 80 g/week increase in legume consumption without changing other elements of the diet. This analysis was also conducted among consumers of legumes only and followed the adjustment levels of model 2 but without excluding any single food components from β3’ and without adjusting for total intake of all foods β2.

To evaluate the robustness of the main analyses, multiple sensitivity analyses were conducted: i) Inclusion of fresh peas in the estimated weekly legume consumption, as fresh peas are a legume pod though traditionally counted as a vegetable in NHS’ 5 A Day recommendations for fruit and vegetables.(^31^) The amount of peas consumed was estimated based on the number of reported portions consumed with one portion of peas weighing 80 g/day.(^31^) ii) Exclusion of soy milk from the estimated weekly legume consumption, as soy milk is unlikely to culinarily replace red and processed meat, poultry, or fish. iii) Excluding participants above the 90^th^ percentile of alcohol intake to reduce the likelihood that cases of fatty liver disease were caused by alcohol intake. iv) Excluding participants with elevated ALT levels cut-off at 40 U/L as defined by guidelines from the European Association for the Study of the Liver^(34; 35)^. v) Excluding participants with < 3 completed Oxford WebQs.

All secondary and sensitivity analyses followed the adjustment level in model 2 from the main analysis (i.e. all covariates were included except BMI).

All analyses were conducted in R (Version 4.1.1, The R Foundation for Statistical Computing) with a two-sided significance level of 5%. The statistical analyses followed a published protocol.(^36^) The statistical procedures only had minor deviations from the protocol. Of note, the non-specific substitution was omitted and replaced by the non-substitution analyses among consumers of legumes. An additional sensitivity analysis excluding soymilk from the legume component was conducted, as replacing animal-based solid foods for a beverage may not be feasible. The statistical analyses were structured to be reproducible using the targets R package(^37^) and all code is available online at https://github.com/steno-aarhus/leha/ (Accessed on October 21, 2024).

## Results

Of the 502,369 participants in the UK Biobank cohort, 126,812 individuals had completed two or more Oxford WebQs. Of these, 2381 were excluded due to missing information on covariates, 192 had an incident event before the start of follow-up, and 45 were lost to follow-up before baseline. This resulted in a study sample of 124,194 participants (54,921 men, 69,273 women) in the analyses (Figure 1). Participants contributed a total of 1,288,233 person-years of follow-up and 1201 individuals developed NAFLD after a median of 10.49 person-years (interquartile range: 10.42-10.92 years).

**Figure 1.**
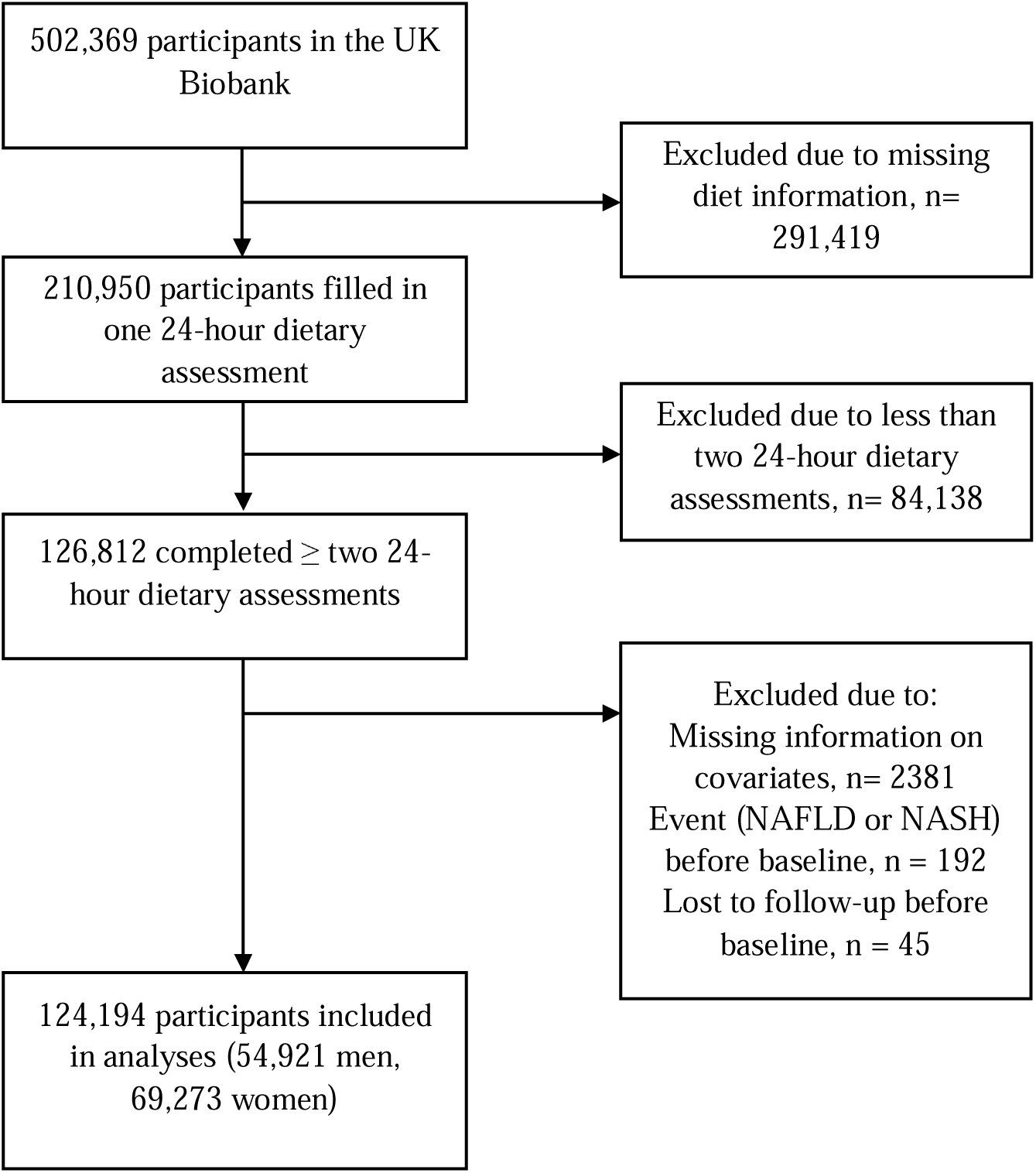
Flowchart of participants in the UK Biobank eligible for inclusion

A large proportion of participants did not report any legume consumption (N = 73,711).

The median (10-90^th^ percentile) age at inclusion was 57.0 (44.0-66.0) years and 56% of participants were women. Individuals with higher legume consumption more often had another ethnicity than White and consumed less alcohol compared to the full cohort. The average weekly intakes of the different food groups were unevenly distributed among those who consumed legumes (across all tertiles) compared to non-consumers. Particularly the consumption of animal-based foods like red and processed meat, poultry, and fish was lower among those with the highest intake of legumes, compared to the full cohort and to those with lower legume consumption (Table 1).

**Table 1.**
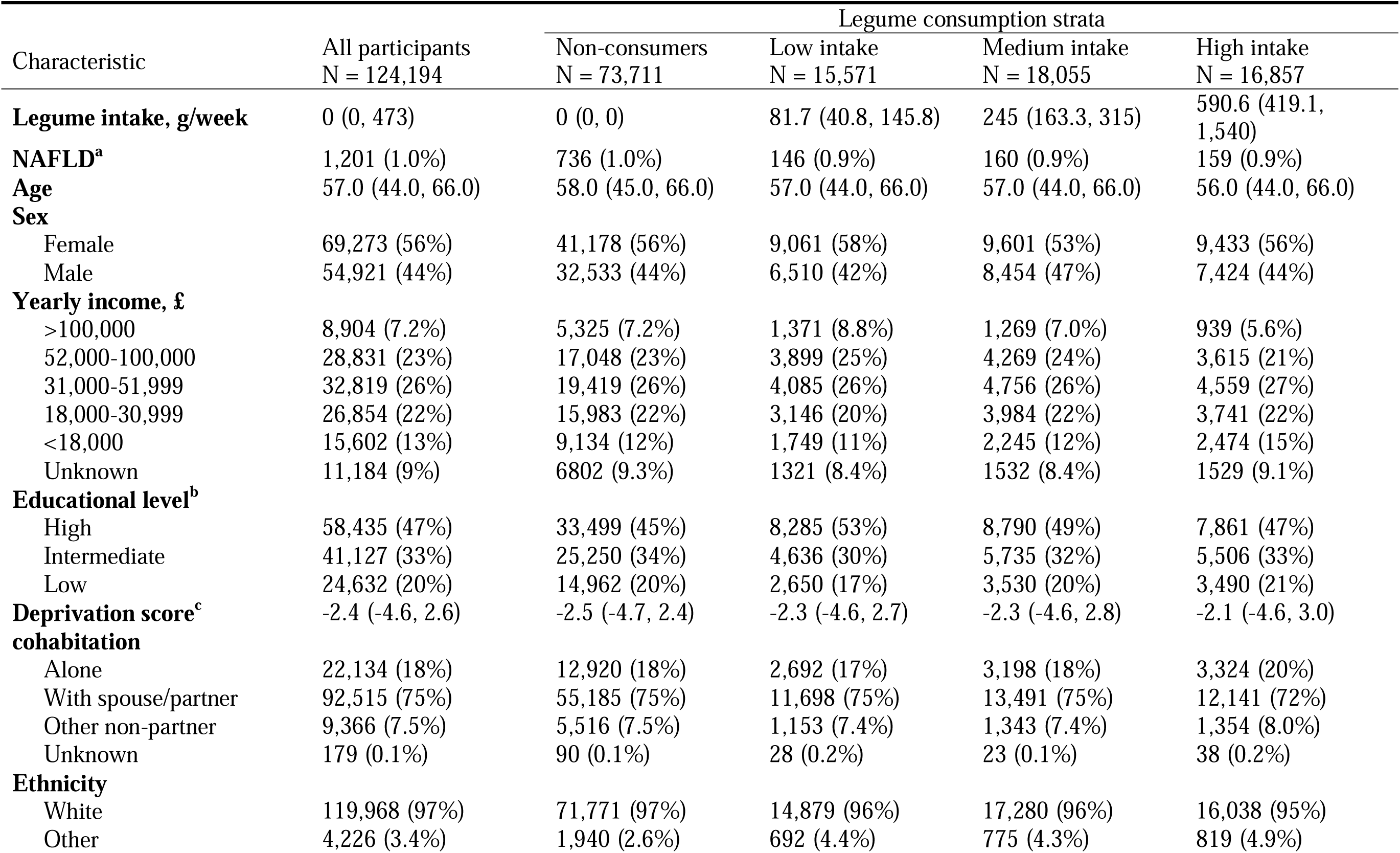
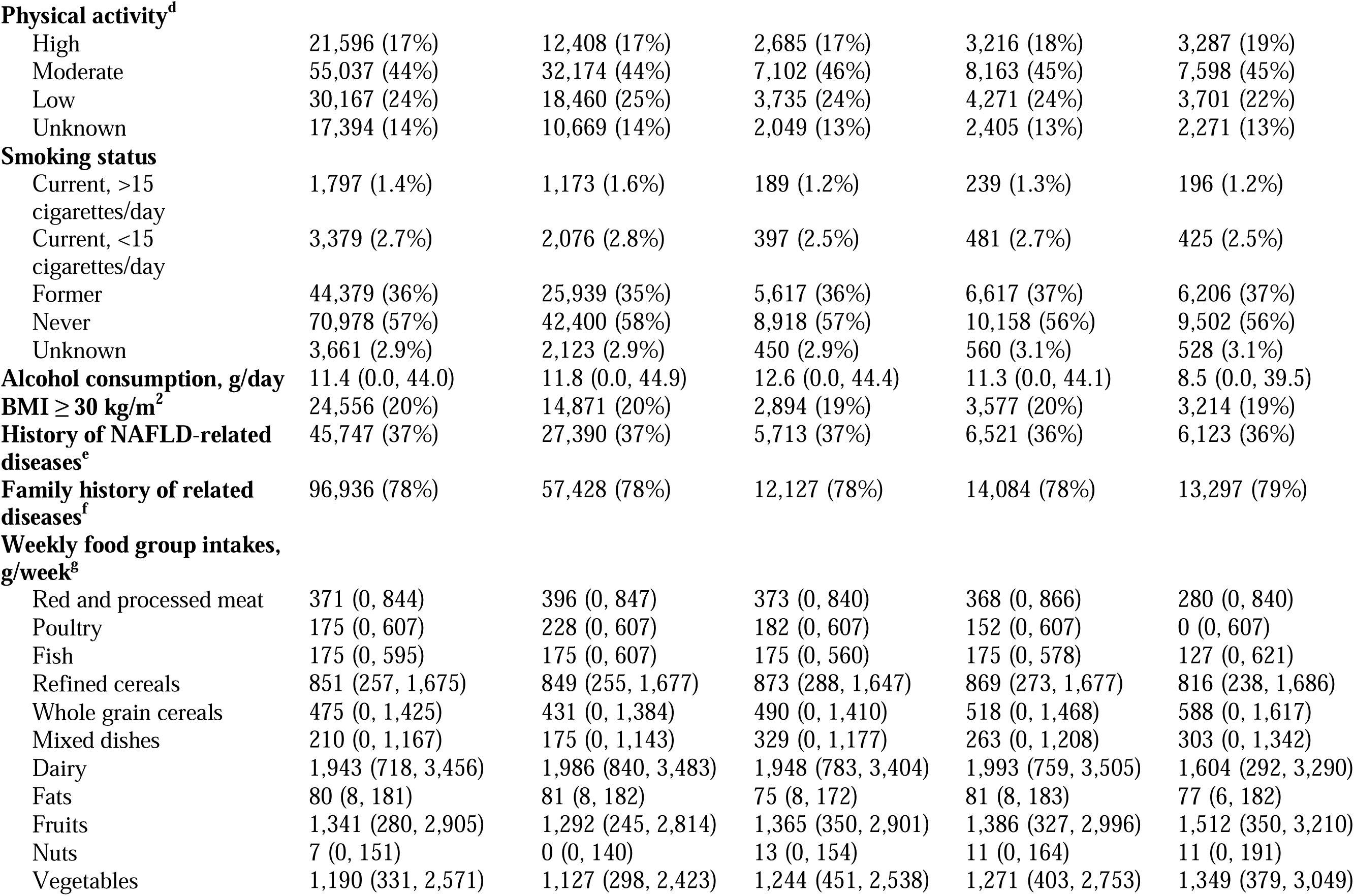
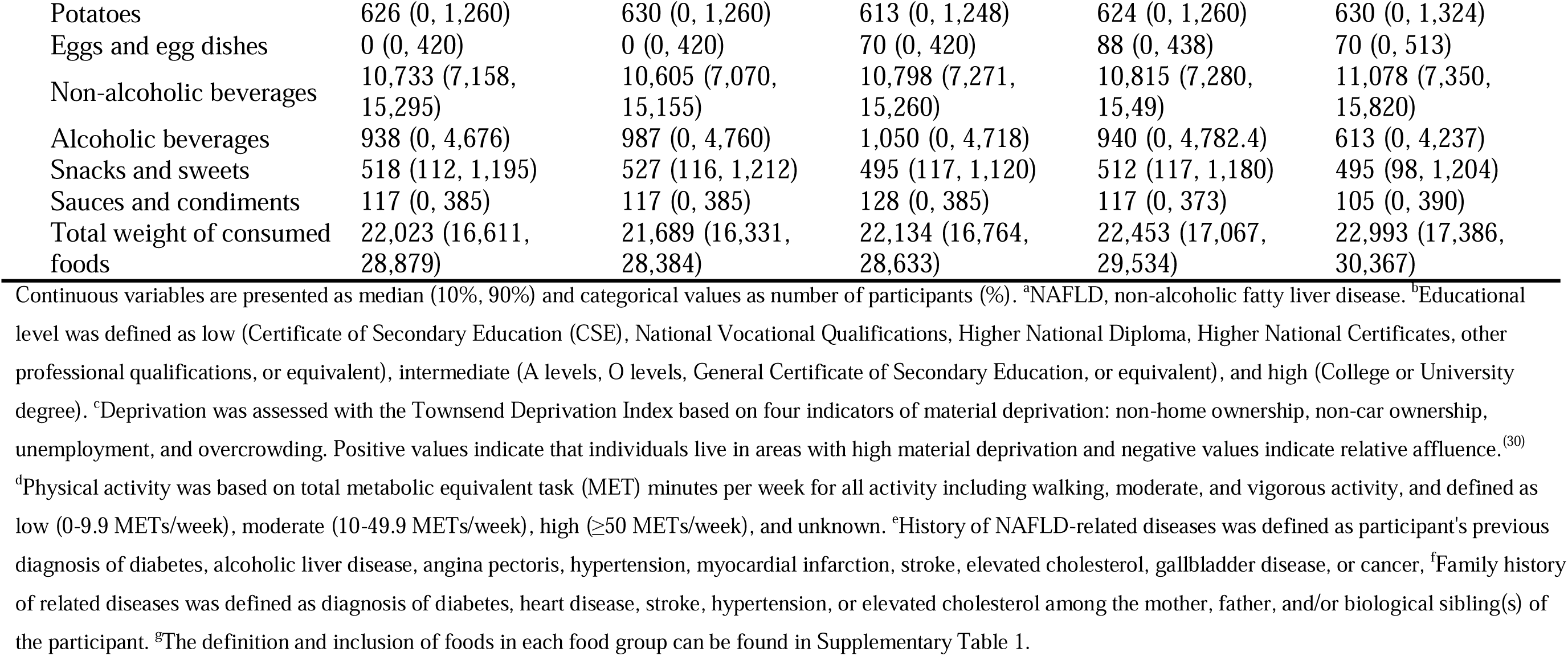
Baseline characteristics across consumption of legumes in the UK Biobank cohort across legume consumption strata (N = 124,194)

| Characteristic | All participants<br>N = 124,194 | Legume consumption strata |  |  |  |
| --- | --- | --- | --- | --- | --- |
|  |  | Non-consumers<br>N = 73,711 | Low intake<br>N = 15,571 | Medium intake<br>N = 18,055 | High intake<br>N = 16,857 |
| **Legume intake, g/week** | 0 (0, 473) | 0 (0, 0) | 81.7 (40.8, 145.8) | 245 (163.3, 315) | 590.6 (419.1, 1,540) |
| **NAFLD<sup>a</sup>** | 1,201 (1.0%) | 736 (1.0%) | 146 (0.9%) | 160 (0.9%) | 159 (0.9%) |
| **Age** | 57.0 (44.0, 66.0) | 58.0 (45.0, 66.0) | 57.0 (44.0, 66.0) | 57.0 (44.0, 66.0) | 56.0 (44.0, 66.0) |
| **Sex** |  |  |  |  |  |
| Female | 69,273 (56%) | 41,178 (56%) | 9,061 (58%) | 9,601 (53%) | 9,433 (56%) |
| Male | 54,921 (44%) | 32,533 (44%) | 6,510 (42%) | 8,454 (47%) | 7,424 (44%) |
| **Yearly income, £** |  |  |  |  |  |
| >100,000 | 8,904 (7.2%) | 5,325 (7.2%) | 1,371 (8.8%) | 1,269 (7.0%) | 939 (5.6%) |
| 52,000-100,000 | 28,831 (23%) | 17,048 (23%) | 3,899 (25%) | 4,269 (24%) | 3,615 (21%) |
| 31,000-51,999 | 32,819 (26%) | 19,419 (26%) | 4,085 (26%) | 4,756 (26%) | 4,559 (27%) |
| 18,000-30,999 | 26,854 (22%) | 15,983 (22%) | 3,146 (20%) | 3,984 (22%) | 3,741 (22%) |
| <18,000 | 15,602 (13%) | 9,134 (12%) | 1,749 (11%) | 2,245 (12%) | 2,474 (15%) |
| Unknown | 11,184 (9%) | 6802 (9.3%) | 1321 (8.4%) | 1532 (8.4%) | 1529 (9.1%) |
| **Educational level<sup>b</sup>** |  |  |  |  |  |
| High | 58,435 (47%) | 33,499 (45%) | 8,285 (53%) | 8,790 (49%) | 7,861 (47%) |
| Intermediate | 41,127 (33%) | 25,250 (34%) | 4,636 (30%) | 5,735 (32%) | 5,506 (33%) |
| Low | 24,632 (20%) | 14,962 (20%) | 2,650 (17%) | 3,530 (20%) | 3,490 (21%) |
| **Deprivation score<sup>c</sup>** | -2.4 (-4.6, 2.6) | -2.5 (-4.7, 2.4) | -2.3 (-4.6, 2.7) | -2.3 (-4.6, 2.8) | -2.1 (-4.6, 3.0) |
| **cohabitation** |  |  |  |  |  |
| Alone | 22,134 (18%) | 12,920 (18%) | 2,692 (17%) | 3,198 (18%) | 3,324 (20%) |
| With spouse/partner | 92,515 (75%) | 55,185 (75%) | 11,698 (75%) | 13,491 (75%) | 12,141 (72%) |
| Other non-partner | 9,366 (7.5%) | 5,516 (7.5%) | 1,153 (7.4%) | 1,343 (7.4%) | 1,354 (8.0%) |
| Unknown | 179 (0.1%) | 90 (0.1%) | 28 (0.2%) | 23 (0.1%) | 38 (0.2%) |
| **Ethnicity** |  |  |  |  |  |
| White | 119,968 (97%) | 71,771 (97%) | 14,879 (96%) | 17,280 (96%) | 16,038 (95%) |
| Other | 4,226 (3.4%) | 1,940 (2.6%) | 692 (4.4%) | 775 (4.3%) | 819 (4.9%) |
| **Physical activity<sup>d</sup>** |  |  |  |  |  |
| High | 21,596 (17%) | 12,408 (17%) | 2,685 (17%) | 3,216 (18%) | 3,287 (19%) |
| Moderate | 55,037 (44%) | 32,174 (44%) | 7,102 (46%) | 8,163 (45%) | 7,598 (45%) |
| Low | 30,167 (24%) | 18,460 (25%) | 3,735 (24%) | 4,271 (24%) | 3,701 (22%) |
| Unknown | 17,394 (14%) | 10,669 (14%) | 2,049 (13%) | 2,405 (13%) | 2,271 (13%) |
| **Smoking status** |  |  |  |  |  |
| Current, >15 cigarettes/day | 1,797 (1.4%) | 1,173 (1.6%) | 189 (1.2%) | 239 (1.3%) | 196 (1.2%) |
| Current, <15 cigarettes/day | 3,379 (2.7%) | 2,076 (2.8%) | 397 (2.5%) | 481 (2.7%) | 425 (2.5%) |
| Former | 44,379 (36%) | 25,939 (35%) | 5,617 (36%) | 6,617 (37%) | 6,206 (37%) |
| Never | 70,978 (57%) | 42,400 (58%) | 8,918 (57%) | 10,158 (56%) | 9,502 (56%) |
| Unknown | 3,661 (2.9%) | 2,123 (2.9%) | 450 (2.9%) | 560 (3.1%) | 528 (3.1%) |
| **Alcohol consumption, g/day** | 11.4 (0.0, 44.0) | 11.8 (0.0, 44.9) | 12.6 (0.0, 44.4) | 11.3 (0.0, 44.1) | 8.5 (0.0, 39.5) |
| **BMI ≥ 30 kg/m<sup>2</sup>** | 24,556 (20%) | 14,871 (20%) | 2,894 (19%) | 3,577 (20%) | 3,214 (19%) |
| **History of NAFLD-related diseases<sup>e</sup>** | 45,747 (37%) | 27,390 (37%) | 5,713 (37%) | 6,521 (36%) | 6,123 (36%) |
| **Family history of related diseases<sup>f</sup>** | 96,936 (78%) | 57,428 (78%) | 12,127 (78%) | 14,084 (78%) | 13,297 (79%) |
| **Weekly food group intakes, g/week<sup>g</sup>** |  |  |  |  |  |
| Red and processed meat | 371 (0, 844) | 396 (0, 847) | 373 (0, 840) | 368 (0, 866) | 280 (0, 840) |
| Poultry | 175 (0, 607) | 228 (0, 607) | 182 (0, 607) | 152 (0, 607) | 0 (0, 607) |
| Fish | 175 (0, 595) | 175 (0, 607) | 175 (0, 560) | 175 (0, 578) | 127 (0, 621) |
| Refined cereals | 851 (257, 1,675) | 849 (255, 1,677) | 873 (288, 1,647) | 869 (273, 1,677) | 816 (238, 1,686) |
| Whole grain cereals | 475 (0, 1,425) | 431 (0, 1,384) | 490 (0, 1,410) | 518 (0, 1,468) | 588 (0, 1,617) |
| Mixed dishes | 210 (0, 1,167) | 175 (0, 1,143) | 329 (0, 1,177) | 263 (0, 1,208) | 303 (0, 1,342) |
| Dairy | 1,943 (718, 3,456) | 1,986 (840, 3,483) | 1,948 (783, 3,404) | 1,993 (759, 3,505) | 1,604 (292, 3,290) |
| Fats | 80 (8, 181) | 81 (8, 182) | 75 (8, 172) | 81 (8, 183) | 77 (6, 182) |
| Fruits | 1,341 (280, 2,905) | 1,292 (245, 2,814) | 1,365 (350, 2,901) | 1,386 (327, 2,996) | 1,512 (350, 3,210) |
| Nuts | 7 (0, 151) | 0 (0, 140) | 13 (0, 154) | 11 (0, 164) | 11 (0, 191) |
| Vegetables | 1,190 (331, 2,571) | 1,127 (298, 2,423) | 1,244 (451, 2,538) | 1,271 (403, 2,753) | 1,349 (379, 3,049) |
| Potatoes | 626 (0, 1,260) | 630 (0, 1,260) | 613 (0, 1,248) | 624 (0, 1,260) | 630 (0, 1,324) |
| Eggs and egg dishes | 0 (0, 420) | 0 (0, 420) | 70 (0, 420) | 88 (0, 438) | 70 (0, 513) |
| Non-alcoholic beverages | 10,733 (7,158, 15,295) | 10,605 (7,070, 15,155) | 10,798 (7,271, 15,260) | 10,815 (7,280, 15,49) | 11,078 (7,350, 15,820) |
| Alcoholic beverages | 938 (0, 4,676) | 987 (0, 4,760) | 1,050 (0, 4,718) | 940 (0, 4,782.4) | 613 (0, 4,237) |
| Snacks and sweets | 518 (112, 1,195) | 527 (116, 1,212) | 495 (117, 1,120) | 512 (117, 1,180) | 495 (98, 1,204) |
| Sauces and condiments | 117 (0, 385) | 117 (0, 385) | 128 (0, 385) | 117 (0, 373) | 105 (0, 390) |
| Total weight of consumed foods | 22,023 (16,611, 28,879) | 21,689 (16,331, 28,384) | 22,134 (16,764, 28,633) | 22,453 (17,067, 29,534) | 22,993 (17,386, 30,367) |

Continuous variables are presented as median (10%, 90%) and categorical values as number of participants (%). <sup>a</sup>NAFLD, non-alcoholic fatty liver disease. <sup>b</sup>Educational level was defined as low (Certificate of Secondary Education (CSE), National Vocational Qualifications, Higher National Diploma, Higher National Certificates, other professional qualifications, or equivalent), intermediate (A levels, O levels, General Certificate of Secondary Education, or equivalent), and high (College or University degree). <sup>c</sup>Deprivation was assessed with the Townsend Deprivation Index based on four indicators of material deprivation: non-home ownership, non-car ownership, unemployment, and overcrowding. Positive values indicate that individuals live in areas with high material deprivation and negative values indicate relative affluence.<sup>(30)</sup>
<sup>d</sup>Physical activity was based on total metabolic equivalent task (MET) minutes per week for all activity including walking, moderate, and vigorous activity, and defined as low (0-9.9 METs/week), moderate (10-49.9 METs/week), high ( $\geq 50$ METs/week), and unknown. <sup>e</sup>History of NAFLD-related diseases was defined as participant's previous diagnosis of diabetes, alcoholic liver disease, angina pectoris, hypertension, myocardial infarction, stroke, elevated cholesterol, gallbladder disease, or cancer, <sup>f</sup>Family history of related diseases was defined as diagnosis of diabetes, heart disease, stroke, hypertension, or elevated cholesterol among the mother, father, and/or biological sibling(s) of the participant. <sup>g</sup>The definition and inclusion of foods in each food group can be found in Supplementary Table 1.

When assessing baseline characteristics and food group intakes in relation to the incidence of NAFLD, those who developed NAFLD had a lower deprivation compared to the full cohort. Individuals who developed NAFLD more often smoked and had higher BMI compared to the full cohort. The median daily alcohol intake was, however, lower among those who developed NAFLD. History of NAFLD-related diseases (individual and family-related) was more pronounced among individuals who developed NAFLD during follow-up (Supplementary Table 2).

Replacing 80 g/week of red and processed meat with legumes was associated with a lower rate of developing NAFLD (HR for model 2: 0.96, 95% CI: 0.94; 0.98; Table 2). Adjusting for BMI (Model 3) changed the magnitude of association slightly (HR: 0.97, 95% CI: 0.95; 0.99). Substituting poultry for legumes was associated with a lower rate of NAFLD (HR for model 2: 0.97, 95% CI: 0.95; 0.99; Table 2). However, this association was not statistically significant after adjusting for BMI. Substituting fish for legumes was not associated with rates of NAFLD (Table 2).

**Table 2.** Hazard ratios and 95 % confidence intervals for non-alcoholic fatty liver disease in the UK Biobank when consuming 80 g/week of legumes in specific and non-specific substitutions, and without food substitution.

| Statistical model | Consuming 80 g/week of legumes instead of 80 g/week of |  |  |
| --- | --- | --- | --- |
|  | Red and processed meat<sup>a</sup> | Poultry<sup>b</sup> | Fish<sup>c</sup> |
| Full sample<br>(N = 124,194, events = 1201) |  |  |  |
| Model 1 | 0.96 (0.94; 0.97) | 0.97 (0.95; 0.99) | 0.99 (0.97; 1.01) |
| Model 2 | 0.96 (0.94; 0.98) | 0.97 (0.95; 0.99) | 0.98 (0.96; 1.01) |
| Model 3 | 0.97 (0.95; 0.99) | 0.98 (0.96; 1.00) | 0.99 (0.97; 1.01) |
| Consumers of legumes<sup>d</sup><br>(N = 50,483, events = 465) |  |  |  |
| Model 2 | 0.97 (0.94; 0.99) | 0.98 (0.95; 1.02) | 0.99 (0.95; 1.02) |

Model 1 was adjusted for age at recruitment, geographical region of recruitment, sex, g/week intake of all other dietary components (red and processed meat, poultry, fish, refined cereal, whole grain cereal, fruits, vegetables, potatoes, nuts, dairy, fats, eggs and egg-dishes, mixed dishes, snacks and sweets, sauce and condiments, non-alcoholic beverages, and alcoholic beverages) apart from the food to be substituted, and total intake of all dietary components in g/week. Model 2 was further adjusted for alcohol intake (ethanol g/week), ethnicity, Townsend Deprivation Index, educational level, yearly income, cohabitation, physical activity, smoking status, history of NAFLD-related diseases, family history of NAFLD-related diseases, and cancer. Model 3 was further adjusted for BMI $\geq 30$ kg/m<sup>2</sup>. <sup>a</sup>Red and processed meat included beef, pork, lamb, and other meats including offal, and sausages, bacon, ham, and liver pâté. <sup>b</sup>Poultry included poultry with or without skin and fried poultry with batter or breadcrumbs. <sup>c</sup>Fish included oily fish, white fish, tinned tuna, fried fish with batter or breadcrumbs, and shellfish. <sup>d</sup>Study sample after excluding participants who reported no consumption of legumes at either of the completed dietary assessments.

### Secondary and sensitivity analyses

When excluding non-consumers of legumes and reducing the sample size to 50,483 participants (n events = 465), the magnitude of associations changed minimally compared to the main analyses, in which all participants were included (Table 2; red meat HR: 0.97, 95% CI: 0.94; 0.99; poultry HR: 0.98, 95% CI: 0.95; 1.02; fish HR: 0.99, 95% CI: 0.95; 1.02).

Consuming 80 g/week more legumes without substituting any other foods was not associated with the rate of NAFLD among consumers (HR: 0.99, 95% CI: 0.98; 1.01).

When including peas in the estimated legume consumption, the magnitude and direction of associations remained similar as in the main analyses, albeit with wider CIs (Table 3; meat HR: 0.95, 95% CI: 0.92; 0.99; poultry HR: 0.97, 95% CI: 0.93; 1.01; fish HR: 0.98, 95% CI: 0.94; 1.02). Excluding soy milk from the estimated legume consumption also resulted in similar rates of NAFLD as in the main analyses (meat HR: 0.96, 95% CI: 0.93; 0.98; poultry HR: 0.97, 95% CI: 0.95; 1.00; fish HR: 0.98, 95% CI: 0.96; 1.01).

**Table 3.** Hazard ratio and 95% confidence intervals for NAFLD when substituting 80 g/week of meat, poultry, or fish with 80 g/week of legumes across altered exposure level and exclusion criteria.

| Altered exposure level | Hazard ratios and 95 % confidence intervals |  |  |
| --- | --- | --- | --- |
|  | Red and processed meat<sup>a</sup> | Poultry<sup>b</sup> | Fish<sup>c</sup> |
| Replacing 80 g/week of legumes, including peas, for animal-based foods, N =124,194 (events = 1201) | 0.95 (0.92; 0.99) | 0.97 (0.93; 1.01) | 0.98 (0.94; 1.02) |
| Replacing 80 g/week of legumes, excluding soymilk, for animal-based foods, N =124,194 (events = 1201) | 0.96 (0.93; 0.98) | 0.97 (0.95; 1.00) | 0.98 (0.96; 1.01) |
| Altered exclusion criteria |  |  |  |
| Individuals with alcohol intake below 90<sup>th</sup> percentile, N= 111,774 (events = 1043) | 0.95 (0.94; 0.97) | 0.97 (0.95; 0.99) | 0.98 (0.96; 1.00) |
| Individuals with normal alanine aminotransferase level<sup>d</sup>, N = 108,754 (events = 823) | 0.96 (0.94; 0.98) | 0.98 (0.95; 1.00) | 0.98 (0.96; 1.01) |
| Individuals with $\geq 3$ 24-hour assessments<sup>e</sup>, N = 76,440 (events = 696) | 0.96 (0.93; 0.98) | 0.98 (0.95; 1.01) | 0.98 (0.95; 1.01) |

Adjustment followed the main analyses Model 2 and adjusted for age at recruitment, geographical region of recruitment, sex, g/week intake of all other dietary components (meat, poultry, fish, refined cereal, whole grain cereal, fruits, vegetables, potatoes, nuts, dairy, fats, eggs and egg-dishes, mixed dishes, snacks and sweets, sauce and condiments, non-alcoholic beverages, and alcoholic beverages) apart from the food to be substituted, total intake of all dietary components in g/week, alcohol intake (ethanol g/week), ethnicity, Townsend Deprivation Index, educational level, yearly income, cohabitation, physical activity, smoking status, history of NAFLD-related diseases, family history of NAFLD-related diseases, and cancer. <sup>a</sup>Red and processed meat included beef, pork, lamb, and other meats including offal, and sausages, bacon, ham, and liver pâté. <sup>b</sup>Poultry included poultry with or without skin and fried poultry with batter or breadcrumbs. <sup>c</sup>Fish included oily fish, white fish, tinned tuna, fried fish with batter or breadcrumbs, and shellfish. <sup>d</sup>Analyses including individuals with alanine aminotransferase levels below 40 U/L. <sup>e</sup>Analyses restricted to individuals with three or more completed 24-hour dietary assessments.

When removing participants with very high alcohol intake or very high ALT circulating concentrations the magnitude and direction of associations remained almost identical to those estimated in main analyses. Exclusion of participants with fewer than three returned Oxford WebQs resulted in marginally widened CIs (Table 3).

## Discussion

In this prospective study of British individuals, we found that replacing 80 g/week of red and processed meat or poultry with legumes was associated with a modestly lower NAFLD rate.

There is still uncertainty about the optimal type of dietary intervention for the prevention of NAFLD, and studies have investigated foods as well as sources of protein. Prospective and cross-sectional studies in humans have indicated that high consumption of animal-based protein particularly from red and processed meat may increase risk of NAFLD, while plant-based proteins appear beneficial.^(3; 38-40)^ To our knowledge, very few substitution studies, replacing animal-based foods for legumes, have investigated the risk of NAFLD. One previous study using two Chinese cohorts investigated replacement of 25 g/day of animal-sourced foods with legumes in relation to NAFLD-rate. They found that replacing poultry with legumes was associated with a lower HR for NAFLD in the Guangzhou Nutrition and Health Study (HR: 0.35, 95%CI: 0.18; 0.69), while replacing red and processed meat or fish with legumes resulted in higher but non-significant rates of NAFLD (meat HR: 1.16, 95% CI: 0.99; 1.37; fish HR: 1.09, 95% CI: 0.92; 1.30). In the Tianjin Chronic Low-grade Systemic Inflammation and Health cohort, replacing processed meat and poultry with legumes was associated with a lower rate of NAFLD, while unprocessed red meat and fish was not associated with NAFLD.(^19^) An Iranian cross-sectional study did not find any association between substituting protein sources (animal-based or plant-based) and NAFLD.(^41^) One study using the UKB substituted 15 g/day of red and processed meat for legumes and found no association with liver cancer, despite this cancer being highly associated with NAFLD. However, these findings could be due to few cases of liver cancer.(^42^)

The substitution effect is a result of including one food and concurrently excluding another food, and it is therefore not possible to assess whether any beneficial association is caused by the higher intake of one food group, or the lower intake of the other food group. By replacing animal-based protein foods like red and processed meat with legumes, intakes of dietary fiber, unsaturated fats, minerals, antioxidants, and phytochemicals are increased. These components may have favorable health impacts and could be key factors contributing to metabolic health, potentially lowering the risk of NAFLD.(^43–46^) In the case of red and processed meat, this substitution may furthermore lower the intakes of saturated fats, cholesterol, heme iron, and daily acid loads. These components may trigger oxidative reactions and pro-inflammatory cytokines, which up-regulate inflammatory responses and potentially increase the risk of developing NAFLD.^(38; 47; 48)^

A substitution size of one weekly serving has previously been argued as a realistic substitution for the general population.(^19^) Such substitutions may also be feasible for dietary recommendations for general populations due to the simple consumer message.(^20^) For a disease like NAFLD with no current medical treatment, it is relevant to identify the optimal diet for preventing disease onset or worsening, and substitution analyses can provide invaluable information by specifying comparisons between individual foods.(^49^) Our results indicate a modest 3-4% lower incidence rate of NAFLD when replacing a serving of red and processed meat or poultry with legumes, but this was not the case for fish.

Compared to research on other foods, very few studies with Western populations have investigated legume intake specifically. This may be due to a negligible legume consumption among many cohorts.^(7; 50-54)^ Other prospective studies, such as the French Etude NutriNet-Santé and the multinational Prospective Urban Rural Epidemiology (PURE) study, also have large proportions of non-consumers of legumes or very low intakes.^(14; 55)^ Large variability in legume consumption exists globally and according to the Global Dietary Database, European countries represent the lowest consumption worldwide as the population in more than one third of all countries consume less than 10 g/day.(^7^) It may also be that weak or null associations in combination with low intakes and high proportions of non-consumers, have led to less focus to publish studies in this research area.^(51-54; 56)^ To address the high proportion of non-consumers in our study population, we excluded non-consumers in sensitivity analyses. The results did not change markedly, but this exclusion limits the generalizability of these results to the general population.

Our substitution models did not adjust for energy intake leading to a caloric difference in the substitution, which would be related to weight and NAFLD (Supplementary Figure 1). Although BMI is clinically on the causal path between food intake and NAFLD, we observed only minor differences in results of our food substitution models with and without adjustment for baseline BMI. We were thus unable to confirm any mediation of the association between replacing red and processed meats, poultry, or fish with legumes through BMI. Similarly, the study on the two Chinese cohorts found mixed associations despite adjusting for BMI.(^19^) Formal causal mediation studies of the role of BMI after baseline on these associations in other studies may provide clearer answers.

This study has several strengths including the prospective cohort design, large sample size and detailed assessment of lifestyle. Furthermore, the food-level substitution analysis is one of the first to investigate the impacts of replacing red and processed meat, poultry, and fish with legumes on the rate of NAFLD. The study does however also have some limitations.

Participants in health research often differentiate from the underlying population on certain characteristics such as health behaviour and literacy. This may introduce selection on the exposure level, where individuals with a higher consumption of healthy foods, such as legumes, are more prone to participate. Our study may therefore not provide accurate estimates of the prevalence of legume intake in the UK due to healthy volunteer bias.

Despite being developed and validated against recovery biomarkers, short-term dietary assessments methods such as the Oxford WebQ have some shortcomings with assessing habitual dietary intake compared to a food frequency questionnaire (FFQ), particularly capturing usual intake of foods that are not consumed very frequently. If diet is only assessed at one time point, the FFQ captures habitual diet better.^(57; 58)^ Including only participants who had completed at least two WebQs ensured more accurate estimation of usual intake compared to relying on a single measurement.^(59; 60)^ This approach also allowed us to adequately capture the range of exposure necessary to examine the association between legume intake and NAFLD. Most participants completed two diet assessments, but usual intake of foods not consumed daily in this population, such as legumes and fish, may not have been fully captured. However, the sensitivity analyses using three dietary assessments did not show a change in magnitude or direction of associations. All participants completed a touchscreen dietary questionnaire at recruitment, which may capture the usual intake of meat and fish intakes better than the 24-hour dietary assessments. However, as legume consumption was not reported separately from vegetables in the touchscreen questionnaire, we were unable to use these data. Furthermore, the prospective study design ensures that participation is non-differential with regards to the outcome, and thus selection bias is unlikely to have affected our results. Despite the thorough adjustment for confounding, residual confounding cannot be ruled out in observational studies. The substitution analyses were furthermore purely observational limiting any causal inference from the results. Future randomized controlled trials would be needed to confirm the findings of our study in a causal setting.

## Conclusion

Replacing red and processed meat or poultry with legumes was associated with a slightly lower rate of NAFLD in this population from the UK. No association was observed when substituting fish for legumes. While the associations identified in this study were modest, our findings align with general recommendations promoting increased consumption of legumes. Further research in populations with higher legume intake is warranted to confirm these findings.

## Acknowledgments

The authors acknowledge the participants who provided data and the members of the UK Biobank cohort who collected the data.

## Transparency Declaration

The lead author affirms that this manuscript is an honest, accurate, and transparent account of the study being reported. The reporting of this work is compliant with the Strengthening the Reporting of Observational Studies in Epidemiology guidelines for nutritional epidemiology (STROBE-nut). The lead author affirms that no important aspects of the study have been omitted and that any discrepancies from the study as planned (https://doi.org/10.5281/zenodo.11670547) have been explained.

## Conflicts of interest

None of the authors have any conflicts of interest to declare.

## Funding

This study was funded by Aarhus University and the Steno Diabetes Center Aarhus. The Graduate School of Health, Aarhus University funded the salary of Fie Langmann. Access to data and data management was funded by the Steno Diabetes Center Aarhus. Daniel B. Ibsen was funded by the Independent Research Fund Denmark with grant number 1057-00016B and the Danish Diabetes Association. The funding agencies had no role in the design and conduct of the study; the collection, management, analysis, and interpretation of the data; the preparation, review, and approval of the manuscript, or the decision to publish the manuscript.

## Data availability

The study was based on data from the UK Biobank prospective cohort that are not publicly available due to personal information. Access to data can be acquired through an application to the Access Management System of UK Biobank online (https://www.ukbiobank.ac.uk/enable-your-research/apply-for-access).

## Notes

### Competing Interest Statement

The authors have declared no competing interest.

### Clinical Protocols

https://doi.org/10.5281/zenodo.11670547

### Author Declarations

The National Information Governance Board for Health and Social Care and the National Health Service (NHS) North West Multicentre Research Ethics Committee (ref 21/NW/0157) gave ethical approval for this work. This work has been conducted using the UK Biobank Resource under Application Number 81520.

## References

1. Wang RZ, Zhang WS, Jiang CQ et al. Association of fish and meat consumption with non-alcoholic fatty liver disease: Guangzhou Biobank Cohort Study. BMC Public Health, 2023; 23: 2433.

2. Tan LJ, Shin S. Effects of oily fish and its fatty acid intake on non-alcoholic fatty liver disease development among South Korean adults. Front Nutr, 2022; 9: 876909.

3. Hashemian M, Merat S, Poustchi H et al. Red Meat Consumption and Risk of Nonalcoholic Fatty Liver Disease in a Population With Low Meat Consumption: The Golestan Cohort Study. Am J Gastroenterol, 2021; 116: 1667–1675.

4. Al-Dayyat HM, Rayyan YM, Tayyem RF. Non-alcoholic fatty liver disease and associated dietary and lifestyle risk factors. Diabetes Metab Synd, 2018; 12: 569–575.

5. Wattacheril JJ, Abdelmalek MF, Lim JK et al. AGA Clinical Practice Update on the Role of Noninvasive Biomarkers in the Evaluation and Management of Nonalcoholic Fatty Liver Disease: Expert Review. Gastroenterology, 2023; 165: 1080–1088.

6. European Association for the Study of the Liver (EASL), European Association for the Study of Diabetes (EASD), European Association for the Study of Obesity (EASO). EASL-EASD-EASO Clinical Practice Guidelines for the management of non-alcoholic fatty liver disease. J Hepatol, 2016; 64: 1388-1402.

7. Yanni AE, Iakovidi S, Vasilikopoulou E et al. Legumes: A Vehicle for Transition to Sustainability. Nutrients, 2023; 16: 98.

8. 8. Food Research Collaboration (2022) Putting climate on everyone’s table: Summary of what the IPCC WG3 report says about food and diet. City University of London.

9. 9. Blomhoff R, Andersen R, Arnesen EK, et al. (2023) Nordic Nutrition Recommendations 2023. https://www.norden.org/en/publication/nordic-nutrition-recommendations-2023

10. Willett W, Rockström J, Loken B et al. Food in the Anthropocene: the EAT-Lancet Commission on healthy diets from sustainable food systems. Lancet, 2019; 393: 447–492

11. Zhao N, Jiao K, Chiu Y-H et al. Pulse Consumption and Health Outcomes: A Scoping Review. Nutrients, 2024; 16: 1435.

12. Rebello CJ, Greenway FL, Finley JW. A review of the nutritional value of legumes and their effects on obesity and its related co-morbidities. Obes Rev, 2014; 15: 392–407.

13. Dinu M, Abbate R, Gensini GF et al. Vegetarian, vegan diets and multiple health outcomes: A systematic review with meta-analysis of observational studies. Crit Rev Food Sci Nutr 2017; 57: 3640–3649.

14. Miller V, Mente A, Dehghan M et al. Fruit, vegetable, and legume intake, and cardiovascular disease and deaths in 18 countries (PURE): a prospective cohort study. Lancet, 2017; 390: 2037–2049.

15. Mudryj AN, Yu N, Hartman TJ et al. Pulse consumption in Canadian adults influences nutrient intakes. Br J Nutr, 2012; 108 S27–36.

16. Hong J, Kim S, Kim H-S. Hepatoprotective Effects of Soybean Embryo by Enhancing Adiponectin-Mediated AMP-Activated Protein Kinase α Pathway in High-Fat and High-Cholesterol Diet-Induced Nonalcoholic Fatty Liver Disease. J Med Food, 2016; 19: 549–559.

17. Son Y, Jang MK, Jung MH. Vigna nakashimae extract prevents hepatic steatosis in obese mice fed high-fat diets. J Med Food, 2014; 17: 1322–1331.

18. Zhang S, Kumari S, Gu Y et al. Soy Food Intake Is Inversely Associated with Newly Diagnosed Nonalcoholic Fatty Liver Disease in the TCLSIH Cohort Study. J Nutr, 2020; 150: 3280–3287.

19. Zhang S, Yan Y, Meng G et al. Protein foods from animal sources and risk of nonalcoholic fatty liver disease in representative cohorts from North and South China. J Intern Med 2023; 293: 340–353.

20. Ibsen DB, Laursen ASD, Würtz AML et al. Food substitution models for nutritional epidemiology. Am J Clin Nutr 2020; 113: 294–303.

21. Song M, Fung TT, Hu FB et al. Association of Animal and Plant Protein Intake With All-Cause and Cause-Specific Mortality. JAMA Intern Med, 2016; 176: 1453–1463.

22. Sudlow C, Gallacher J, Allen N et al. UK biobank: an open access resource for identifying the causes of a wide range of complex diseases of middle and old age. PLoS Med, 2015; 12: e1001779.

23. Watling CZ, Kelly RK, Dunneram Y et al. Associations of intakes of total protein, protein from dairy sources, and dietary calcium with risks of colorectal, breast, and prostate cancer: a prospective analysis in UK Biobank. Br J Cancer, 2023; 129: 636–647.

24. Greenwood DC, Hardie LJ, Frost GS et al. Validation of the Oxford WebQ Online 24-Hour Dietary Questionnaire Using Biomarkers. Am J Epidemiol, 2019; 188: 1858–1867.

25. Perez-Cornago A, Pollard Z, Young H et al. Description of the updated nutrition calculation of the Oxford WebQ questionnaire and comparison with the previous version among 207,144 participants in UK Biobank. Eur J Nutr, 2021; 60: 4019–4030.

26. Piernas C, Perez-Cornago A, Gao M et al. Describing a new food group classification system for UK biobank: analysis of food groups and sources of macro- and micronutrients in 208,200 participants. Eur J Nutr, 2021; 60: 2879–2890.

27. 27. NHS Digital (2021) NHS Data Model and Dictionary International Classification of Diseases (ICD). https://datadictionary.nhs.uk/supporting_information/international_classification_of_diseases__icd_.html (accessed 14 October 2021)

28. Rinella ME, Lazarus JV, Ratziu V et al. A multisociety Delphi consensus statement on new fatty liver disease nomenclature. Hepatology, 2023; 78: 1966–1986.

29. 29. UK Biobank (2020) Hospital inpatient data. UK Biobank.

30. 30. National Centre for Research Methods (n.d.) Geographical Referencing Learning Resources - Townsend Deprivation Index. https://www.restore.ac.uk/geo-refer/36229dtuks00y19810000.php (accessed 23 April 2024)

31. 31. UK National Health Services (2022) 5 A Day portion sizes. https://www.nhs.uk/live-well/eat-well/5-a-day/portion-sizes/ (accessed 16 December 2022)

32. Tomova GD, Arnold KF, Gilthorpe MS et al. Adjustment for energy intake in nutritional research: a causal inference perspective. Am J Clin Nutr, 2022; 115: 189–198.

33. Tomova GD, Gilthorpe MS, Tennant PWG. Theory and performance of substitution models for estimating relative causal effects in nutritional epidemiology. The American Journal of Clinical Nutrition, 2022; 116: 1379–1388.

34. Radonjić T, Milićević O, Jovanović I et al. Elevated Transaminases as Predictors of COVID-19 Pneumonia Severity. Medicina (Kaunas), 2022; 58.

35. Claus M, Antoni C, Hofmann B. Factors associated with elevated alanine aminotransferase in employees of a German chemical company: results of a large cross-sectional study. BMC Gastroenterol, 2021; 21: 25.

36. Langmann F, Ibsen DB, Johnston LW, et al. Protocol: Substituting red and processed meat, poultry, and fish with legumes and risk of non-alcoholic fatty liver disease in a large prospective cohort. Zenodo, 2024.

37. Landau W. The targets R package: a dynamic Make-like function-oriented pipeline toolkit for reproducibility and high-performance computing. Journal of Open Source Software, 2021; 6: 2959.

38. Alferink LJ, Kiefte-de Jong JC, Erler NS et al. Association of dietary macronutrient composition and non-alcoholic fatty liver disease in an ageing population: the Rotterdam Study. Gut, 2019; 68: 1088–1098.

39. Noureddin M, Zelber-Sagi S, Wilkens LR et al. Diet Associations With Nonalcoholic Fatty Liver Disease in an Ethnically Diverse Population: The Multiethnic Cohort. Hepatology, 2020; 71: 1940–1952.

40. Rietman A, Sluik D, Feskens EJM et al. Associations between dietary factors and markers of NAFLD in a general Dutch adult population. Eur J Clin Nutr, 2018; 72: 117–123.

41. Amirkalali B, Khoonsari M, Sohrabi MR et al. Relationship between dietary macronutrient composition and non-alcoholic fatty liver disease in lean and non-lean populations: a cross-sectional study. Public Health Nutr 2021; 24: 6178–6190.

42. Bock N, Langmann F, Johnston LW et al. The Association between the Substitution of Red Meat with Legumes and the Risk of Primary Liver Cancer in the UK Biobank: A Cohort Study. Nutrients, 2024; 16: 2383.

43. Satija A, Bhupathiraju SN, Spiegelman D et al. Healthful and Unhealthful Plant-Based Diets and the Risk of Coronary Heart Disease in U.S. Adults. J Am Coll Cardiol, 2017; 70: 411–422.

44. Ferreira H, Vasconcelos M, Gil AM et al. Benefits of pulse consumption on metabolism and health: A systematic review of randomized controlled trials. Crit Rev Food Sci Nutr, 2021; 61: 85–96.

45. Bouchenak M, Lamri-Senhadji M. Nutritional quality of legumes, and their role in cardiometabolic risk prevention: a review. J Med Food, 2013; 16: 185–198.

46. Mudryj AN, Yu N, Aukema HM. Nutritional and health benefits of pulses. Appl Physiol Nutr Metab, 2014; 39: 1197–1204.

47. Ma J, Hennein R, Liu C et al. Improved Diet Quality Associates With Reduction in Liver Fat, Particularly in Individuals With High Genetic Risk Scores for Nonalcoholic Fatty Liver Disease. Gastroenterology, 2018; 155: 107–117.

48. Sivasubramanian BP, Dave M, Panchal V et al. Comprehensive Review of Red Meat Consumption and the Risk of Cancer. Cureus, 2023; 15: e45324.

49. Song M, Giovannucci E. Substitution analysis in nutritional epidemiology: proceed with caution. Eur J Epidemiol, 2018; 33: 137–140.

50. Faber I, Henn K, Brugarolas M et al. Relevant characteristics of food products based on alternative proteins according to European consumers. J Sci Food Agric, 2021.

51. Mertens E, Kuijsten A, Dofková M et al. Geographic and socioeconomic diversity of food and nutrient intakes: a comparison of four European countries. Eur J Nutr, 2019; 58: 1475–1493.

52. Rawal V, Charrondiere R, Xipsiti M et al. (2019) The Global Economy of Pulses. Food and Agriculture Organization of the United Nations.

53. Figueira N, Curtain F, Beck E et al. Consumer Understanding and Culinary Use of Legumes in Australia. Nutrients, 2019; 11: 1575.

54. Perera T, Russo C, Takata Y et al. Legume Consumption Patterns in US Adults: National Health and Nutrition Examination Survey (NHANES) 2011–2014 and Beans, Lentils, Peas (BLP) 2017 Survey. Nutrients, 2020; 12: 1237.

55. Berthy F, Brunin J, Allès B et al. Association between adherence to the EAT-Lancet diet and risk of cancer and cardiovascular outcomes in the prospective NutriNet-Santé cohort. The American Journal of Clinical Nutrition, 2022; 116: 980–991.

56. Sharma H, Verma S. Is positive publication bias really a bias, or an intentionally created discrimination toward negative results? Saudi J Anaesth, 2019; 13: 352–355.

57. Freedman LS, Midthune D, Arab L et al. Combining a Food Frequency Questionnaire With 24- Hour Recalls to Increase the Precision of Estimation of Usual Dietary Intakes-Evidence From the Validation Studies Pooling Project. Am J Epidemiol, 2018; 187: 2227–2232.

58. Park Y, Dodd KW, Kipnis V et al. Comparison of self-reported dietary intakes from the Automated Self-Administered 24-h recall, 4-d food records, and food-frequency questionnaires against recovery biomarkers. The American Journal of Clinical Nutrition, 2018; 107: 80-93.

59. Carter JL, Lewington S, Piernas C et al. Reproducibility of dietary intakes of macronutrients, specific food groups, and dietary patterns in 211 050 adults in the UK Biobank study. J Nutr Sci, 2019; 8: e34.

60. Galante J, Adamska L, Young A et al. The acceptability of repeat Internet-based hybrid diet assessment of previous 24-h dietary intake: administration of the Oxford WebQ in UK Biobank. British Journal of Nutrition, 2016; 115: 681–686.

