## Supplementary information for "Legumes as a substitute for red and processed meat, poultry, or fish, and the risk of non-alcoholic fatty liver disease in a large cohort"

Supplementary Table 1. Description of dietary components included in the definition of food groups for the study.

| Food group | Dietary components |
|---|---|
| Legumes | Legumes and pulses, soy desserts and yoghurt, soy milk, meat substitutes made from soy. |
| Red and processed meat | Beef, pork, lamb, offal, sausages, bacon, ham, and liver pâté. |
| Poultry | Poultry with or without skin and fried poultry with batter or breadcrumbs. |
| Fish | Oily fish, white fish, tinned tuna, fried fish with batter or breadcrumbs, and shellfish. |
| Eggs and egg dishes | Eggs and egg-based dishes like scrambled eggs, omelets, and quiche. |
| Refined cereal | White pasta and rice, white bread, biscuit cereal, biscuits, savory crackers, mixed bread (50/50 brown and seeded), other bread, and other cereal with high sugar content. |
| Whole grain cereal | Whole meal bread, whole meal pasta, brown rice, bran cereal, oat cereal (with and without added sugar), muesli, and other whole grains. |
| Dairy | Milk (whole, semi-skimmed, skimmed), cream, cheese (low, medium, and high fat), yoghurt (low or full fat), milk-based and powdered drinks, and milk-dairy desserts. |
| Fruits | Berries, citrus fruits, apples, pears, dried fruits, stewed fruits, and other fruits. |
| Vegetables | Green leafy vegetables, cabbages, onion, garlic, root vegetables, raw salad, peas, sweetcorn, vegetable dips (guacamole and hummus), tomatoes, mushrooms, mixed vegetables, and vegetable side dishes. |
| Potatoes | Fried/roasted potatoes, mashed potatoes, and baked or boiled potatoes and sweet potatoes. |
| Nuts | Salted and unsalted nuts and seeds. |
| Fats | Olive oil, animal-based fat spreads, and plant-based fat spreads. |

| | |
|---|---|
| Mixed dishes | Pizza, grain dishes with added fat, samosa, pakora, soups, sushi, and vegetarian non-soy meat substitutes like quorn. |
| Sauce and condiments | Sauces and condiments with low or high fat content. |
| Snacks and sweets | Nut-based spreads, savoury snacks, chocolate confectionery, added sugars and preserves, cakes, pastry, other desserts, and other sweets. |
| Non-alcoholic beverages | Coffee, tea, rice milk, oat milk, water (still or sparkling), sugar-sweetened soft drinks, artificially sweetened soft drinks, and fruit juices. |
| Alcoholic beverages | Beer, cider, spirits, and fortified and non-fortified wines. |

Supplementary Table 2. Baseline characteristics of participants in the UK Biobank cohort across incident NAFLD (N = 123,586)

| **Characteristic** | **All participants**<br>N = 124,194 | **With NAFLD**[a]<br>N = 1201 |
|---|---|---|
| **Age** | 57.0 (44.0, 66.0) | 57.0 (45.6, 66.0) |
| **Sex** | | |
| Female | 69,273 (56%) | 618 (51%) |
| Male | 54,921 (44%) | 583 (49%) |
| **Yearly income, £** | | |
| >100,000 | 8,904 (7.2%) | 54 (4.5%) |
| 52,000-100,000 | 28,831 (23%) | 209 (17%) |
| 31,000-51,999 | 32,819 (26%) | 289 (24%) |
| 18,000-30,999 | 26,854 (22%) | 292 (24%) |
| <18,000 | 15,602 (13%) | 227 (19%) |
| Unknown | 11,184 (9.0%) | 130 (11%) |
| **Educational level**[b] | | |
| High | 58,435 (47%) | 423 (35%) |
| Intermediate | 41,127 (33%) | 442 (37%) |
| Low | 24,632 (20%) | 336 (28%) |
| **Deprivation score**[c] | -2.4 (-4.7, 2.6) | -1.7 (-4.5, 3.6) |
| **Cohabitation** | | |
| Alone | 22,134 (18%) | 241 (20%) |
| With spouse/partner | 92,515 (74%) | 833 (69%) |
| Other non-partner | 9,366 (7.5%) | 121 (10%) |
| Unknown | 179 (0.1%) | 6 (0.5%) |
| **Ethnicity** | | |
| White | 119,968 (97%) | 1,147 (96%) |
| Other | 4,226 (3.4%) | 54 (4.5%) |
| **Physical activity**[d] | | |
| High | 21,596 (17%) | 176 (15%) |
| Moderate | 55,037 (44%) | 469 (39%) |
| Low | 30,167 (24%) | 370 (31%) |
| Unknown | 17,394 (14%) | 186 (15%) |

| | | |
|---|---|---|
| **Smoking status** | | |
| Current, >15 cigarettes/day | 1,797 (1.4%) | 33 (2.7%) |
| Current, <15 cigarettes/day | 3,379 (2.7%) | 45 (3.7%) |
| Former | 44,379 (36%) | 509 (42%) |
| Never | 70,978 (57%) | 570 (47%) |
| Unknown | 3,661 (2.9%) | 44 (3.7%) |
| **Alcohol consumption, g/day** | 11.4 (0.0, 44.0) | 9.0 (0.0, 51.4) |
| **UK-region of recruitment** | | |
| East Midlands | 9,931 (8.0%) | 101 (8.4%) |
| London | 25,432 (20%) | 282 (23%) |
| North East | 11,794 (9.5%) | 156 (13%) |
| North West | 15,291 (12%) | 205 (17%) |
| Scotland | 6,577 (5.3%) | 15 (1.2%) |
| South East | 11,656 (9.4%) | 72 (6.0%) |
| South West | 12,917 (10%) | 122 (10%) |
| Wales | 3,876 (3.1%) | 11 (0.9%) |
| West Midlands | 7,542 (6.1%) | 66 (5.5%) |
| Yorkshire and Humber | 19,178 (15%) | 171 (14%) |
| **BMI ≥ 30 kg/m$^2$** | 24,556 (20%) | 592 (49%) |
| **History of NAFLD-related diseases[e]** | 45,747 (37%) | 715 (60%) |
| **Family history of related diseases[f]** | 96,936 (78%) | 982 (82%) |
| **Food group intakes, g/week[g]** | | |
| Legumes | 0.0 (0.0, 472.5) | 0.0 (0.0, 472.5) |
| Red and processed meat | 371.0 (0.0, 843.5) | 420.0 (0.0, 1,022.0) |
| Poultry | 175.0 (0.0, 606.7) | 227.5 (0.0, 606.7) |
| Fish | 175.0 (0.0, 595.0) | 161.0 (0.0, 581.0) |
| Eggs and egg dishes | 0.0 (0.0, 420.0) | 0.0 (0.0, 513.3) |
| Refined cereal | 850.5 (257.3, 1,674.6) | 855.8 (269.5, 1,659.0) |
| Whole grain cereal | 474.8 (0.0, 1,424.5) | 350.0 (0.0, 1,361.5) |
| Dairy | 1,942.5 (717.5, 3,456.3) | 1,837.5 (600.8, 3,517.5) |
| Fruits | 1,341.2 (280.0, 2,905.0) | 1,190.0 (70.0, 2,968.0) |

| | | |
|---|---|---|
| Vegetables | 1,190.0 (330.8, 2,570.6) | 1,114.8 (236.3, 2,637.3) |
| Potatoes | 625.6 (0.0, 1,260.0) | 630.0 (0.0, 1,365.0) |
| Nuts | 7.0 (0.0, 150.5) | 0.0 (0.0, 140.0) |
| Fats | 79.6 (7.6, 180.9) | 86.9 (8.4, 198.6) |
| Mixed dishes | 210.0 (0.0, 1,166.7) | 210.0 (0.0, 1,166.7) |
| Sauce and condiments | 116.7 (0.0, 385.0) | 105.0 (0.0, 385.0) |
| Snacks and sweets | 518.0 (112.0, 1,194.7) | 556.5 (116.7, 1,354.5) |
| Non-alcoholic beverages | 10,733.3 (7,157.5, 15,295.0) | 10,990.0 (7,093.3, 15,890.0) |
| Alcoholic beverages | 938.0 (0.0, 4,676.0) | 721.9 (0.0, 5,496.8) |
| Total weight of food and beverages | 22,023.0 (16,611.0, 28,878.6) | 22,325.6 (16,421.8, 29,921.8) |

Continuous variables are presented as median (10%, 90%) and categorical values are presented as numbers (%). [a]NAFLD, non-alcoholic fatty liver disease. [b]Educational level was defined as low (Certificate of Secondary Education (CSE), National Vocational Qualifications, Higher National Diploma, Higher National Certificates, other professional qualifications, or equivalent), intermediate (A levels, O levels, General Certificate of Secondary Education, or equivalent), and high (College or University degree). [c]Deprivation was assessed with the Townsend Deprivation Index based on four indicators of material deprivation: non-home ownership, non-car ownership, unemployment, and overcrowding. Positive values indicate that individuals live in areas with high material deprivation and negative values indicate relative affluence.[(84)] [d]Physical activity was based on total metabolic equivalent task (MET) minutes per week for all activity including walking, moderate, and vigorous activity, and defined as low (0-9.9 METs/week), moderate (10-49.9 METs/week), high (≥50 METs/week), and unknown. [e]History of NAFLD-related diseases was defined as participant's previous diagnosis of diabetes, alcoholic liver disease, angina pectoris, hypertension, myocardial infarction, stroke, elevated cholesterol, or gallbladder disease), [f]Family history of related diseases was defined as diagnosis of diabetes, heart disease, stroke, hypertension, or elevated cholesterol among the mother, father, and/or biological sibling(s) of the participant. [g]The definition and inclusion of foods in each food group can be found in Supplementary Table 1.

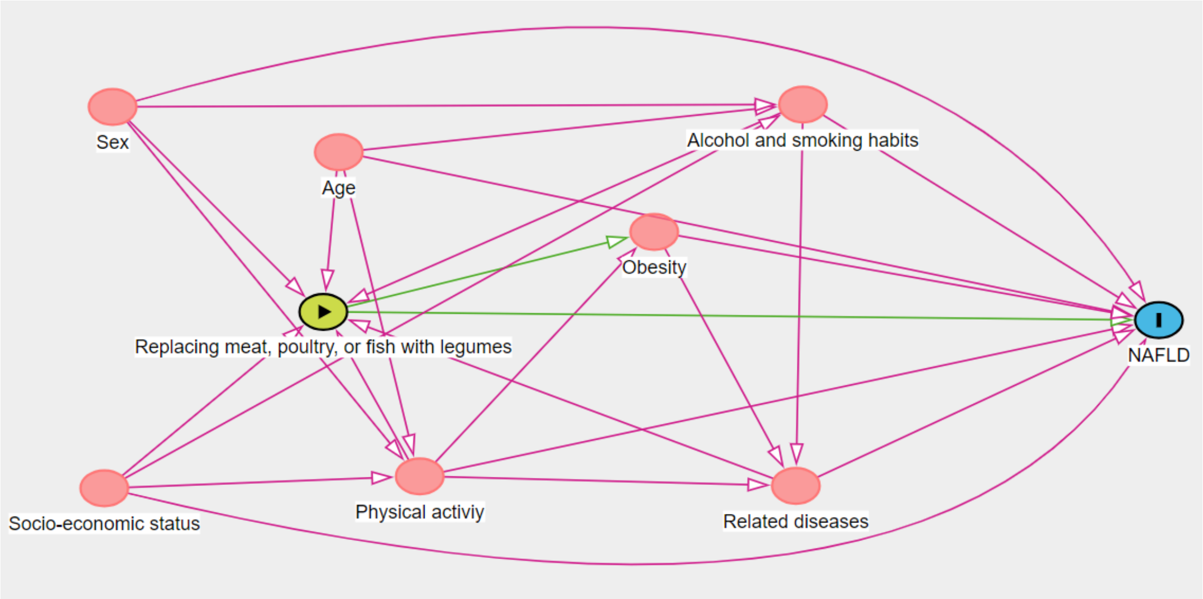

Supplementary Figure 1. Directed Acyclic Graph representing the association between replacing meat, poultry, or fish with legumes and risk of non-alcoholic fatty liver disease (NAFLD) and the assumed relationship with covariates. Socio-economic status comprises factors like ethnicity, Townsend Deprivation Index, geographical living area, cohabitation status, educational level, and yearly income. Related diseases include cancer, hypertension, stroke, myocardial infarction, elevated cholesterol, gallbladder diseases, alcoholic liver disease, diabetes or family history of hypertension, diabetes, stroke, or heart disease. exposure, outcome, ancestor of outcome, ancestor of exposure and outcome, causal path, biasing path
